# Socioeconomic and comorbid factors affecting mortality and length of stay in COVID-19

**DOI:** 10.1101/2021.08.26.21262693

**Authors:** Adam V. Delora, Ashlynn Mills, David Jacobson, Brendon Cornett, W. Frank Peacock, Anita Datta, Shane Jenks

**Affiliations:** HCA Houston Healthcare/University of Houston College of Medicine Consortium; University of Houston College of Medicine; HCA Healthcare; Baylor College of Medicine; HCA Houston Healthcare/University of Houston College of Medicine Consortium, Envision Healthcare

## Abstract

**Background:** The severe acute respiratory syndrome coronavirus 2 (SARS-CoV-2) pandemic exposed and exacerbated health disparities between socioeconomic groups. Our purpose was to determine which disparities are most prevalent and their impact on length of stay (LoS) and in hospital mortality in patients diagnosed with Covid-19.

**Methods:** De-Identified data for patients who tested positive for COVID-19 was abstracted from the HCA enterprise database. Data was binned into summary tables. A negative binomial regression with LoS as the dependent variable and a logistic regression of in-hospital mortality data, using age, insurance status, sex, comorbidities as the dependent variables, were performed.

**Results:** From March 1, 2020 to August 23, 2020, of 111,849 covid testing patient records, excluding those with missing data (n=7), without confirmed COVID-19 (n=27,225), and those from a carceral environment (n=1,861), left 84,624 eligible patients. Compared to the US population, the covid cohort had more black patients (23.17% vs 13.4%). Compared to the white cohort, the black cohort had higher private insurance rates (28.52% vs. 23.68%), shorter LoS (IRR=0.97 CI=0.95-0.99, P<0.01) and lower adjusted mortality (OR 0.81, 95% CI 0.75-0.97). Increasing age was associated with increased mortality and LoS. Patients with Medicare or Medicaid had longer LoS (IRR=1.07, 95% CI=1.04-1.09) and higher adjusted mortality rates (OR=1.11, 95% CI=1-1.23) than those with private insurance

**Conclusion:** Conclusions We found that when blacks have higher rates of private insurance, they have shorter hospitalizations and lower mortality than whites, when diagnosed with Covid-19. Some other psychiatric and medical conditions also significantly impacted outcomes in patients with Covid-19.

**Key Points:** *Question:* Which social determinants of health and comorbidities are most prevalent and their impact on length of stay and in-hospital mortality in patients diagnosed with Covid-19?

*Findings:* In this retrospective cohort of 84,624 with the black cohort having higher private insurance rates (28.52% vs. 23.68%), there was shorter LoS (IRR=0.97 CI=0.95-0.99, P<0.01) and lower adjusted mortality (OR 0.81, 95% CI 0.75-0.97). Age and several other medical and psychiatric comorbidities were also found to correlate with length of stay and mortality.

*Meaning:* The genetics of race is not important in predicting mortality and length of stay in COVID-19 patients, but age, comorbidities, and insurance status appear to have a significant difference in mortality and length of stay.

## Introduction

In December 2019, an outbreak of severe acute respiratory syndrome coronavirus 2 (SARS-CoV-2) occurred and resulted in a global spread.^1^ Several articles have been published looking at health disparities among different populations with COVID-19 and have shown poorer outcomes in minority communities.^2-13^ Early in the pandemic, several papers were published that looked at these disparities and hypothesized factors that would result in a greater risk for infection among Black Americans. Some of these theoretical factors were densely populated housing, a greater burden of chronic disease, limited healthcare access, higher poverty rates, and higher likelihood of employment as essential workers.^14,15^ However, while systemic inequality has been conclusively shown to contribute to morbidity and mortality in other infectious diseases, this connection has not been adequately established with COVID-19 infections.^16-22^ Systemic social inequality and discrepancies in socioeconomic status (SES) contribute to higher incidence of asthma, CVD, hypertension, CKD, and obesity in segments of the general population.^23^ These social disparities point to a national need for patient data analysis to describe and substantiate the various disparities and their associated factors.

Despite a firm understanding on the impact of social determinants on health outcomes, the elemental factors guiding infection stand to be investigated. In Texas, counties with the highest incidence rate (IR) of COVID-19 infection had a higher proportion of black residents, a higher unemployment rate, and a lower proportion of smoking in population as compared to counties with lower IRs.^24^ Counties with higher proportions of black residents have higher prevalence of comorbidities and greater air pollution nationally.^6^

In both persons and populations, Social Determinants of Health (SDoH) may serve a larger impact on health outcomes than inherited components. The social and community context of SDoH classifies individual and structural discrimination as well as social support and social capital as critical components of an individual’s well-being. In data extracted from COVID-NET and National Center for Health Statistics (NCHS), the CDC reports increased hospitalization rates of 2.9 times and 2.8 times more in black Americans and Hispanic patients respectively when compared to white non-Hispanic Americans. They also note a death rate of 1.9 times in Black Americans and a rate of 2.3 times in Hispanic and Latino populations. This data does not consider comorbidities that affect outcomes in COVID-19. The CDC investigated several factors that may affect mortality in COVID-19 including long-standing systemic health and social inequities. Unfortunately, because much of the data is missing in their populations, with 40-50% of race and ethnicity data unaccounted for, strong conclusions about the data cannot be made.^13, 25-30^

Assessing socioeconomic status can be difficult. We propose the use of health insurance types as a proxy. According to the census bureau in 2019, “87.6 percent of people with income-to-poverty ratio at or above 400 percent of poverty had private coverage, compared with 60.1 percent for those with incomes 100 to 399 percent of poverty, and 26.6 percent for those with incomes below poverty”. In contrast, public coverage was the most prevalent for the population in poverty (65.2 percent) and least prevalent for the population with income-to-poverty ratios at or above 400 percent of poverty (21.9 percent). Medicaid requires that an applicant be under a certain income. Also, of note under the affordable care act, states can expand Medicaid eligibility to people whose income-to-poverty ratio falls under a particular threshold, but the correlation still hold even though the relationship between poverty status and health insurance coverage in 2019 may be related to the state of residence and whether that state expanded Medicaid eligibility.^31^

Understanding the effects of SDoH and their interactions with comorbidities can help facilitate rational public programs and may contribute to our knowledge of health disparities in the context of the COVID-19 pandemic and in other medical conditions and in future pandemics. To identify groups who are most likely to have poor outcomes, high-quality data on socioeconomic factors are needed, which will have important implications in the development of public health measures.^32^ HCA is the largest health system in the United States and thus provides an appropriate database to evaluate SDoH in relation to SARS CoV2 infection associated outcomes. Given that HCA is a private hospital system, less demographic differences are expected with fewer gaps in data given that data is more uniformly gathered throughout the network.

This analysis will look at how demographics interact with comorbidities to produce different outcomes in different populations. Understanding the effects of SDoH and their interactions with comorbidities can help facilitate rational public programs and may contribute to our knowledge of health disparities in the context of the COVID-19 pandemic and in other medical conditions and in future pandemics. Our purpose was to determine which disparities are most prevalent and their impact on length of stay (LoS) and in-hospital mortality in patients diagnosed with Covid-19.

## Methods

This is a retrospective observational analysis, using de-identified electronic medical record (EMR) data from 182 HCA hospitals across the US. Patient level data was eligible for inclusion if they tested positive for COVID-19 with PCR or antigen test from March 1, 2020 to August 23, 2020. Patients were excluded from analysis if they did not have Covid-19, for missing data, or if they were transferred from a carceral environment. With in-hospital mortality data as the dependent variable we performed logistic regression with penalized profile likelihood with a pseudo-observation indicator (FLAC model) using age, insurance, sex, comorbidites as the dependent variables. Data is presented with 95% Confidence intervals (95%CI) and odds ratios (OR) and incidence rate ratio (IRR). In both models we also included an interaction term that shows charity insurance and black race vs all others. To account for demographic differences in our population versus the CDC data we also performed a regression model looking at insurance status and race with the outcomes of in-hospital mortality and LoS. Finally, we used the Elixhauser Comorbidity index to define the consequence of multiple concurrent morbidities.^33^

## Results

There were 111,849 patient records initially selected. A total of 27,225 patients were excluded because 25,357 did not have a confirmed covid infection, 7 were missing demographic data (gender, race, etc.) and 1,861 patients were remanded to law enforcement or were transferred from a carceral environment. This population was deemed not representative of the population of interest. The remaining 84,624 patients constitute the analytic cohort.

Our population differs in socioeconomic status by race as well as comorbidities. Demographics and hospital stay characteristics are presented in Table 1. Our population self-identified as 52.38% female, and 49.09% white, 23.17% black, 25.26% multiracial, and 0.22% hispanic. We have a larger black population than in the general population of the United States of America, at 23.17% vs 13.4%. The percentage of our population that considers themselves multiracial or does not fit into the predefined groups by which the data is entered is much larger than the general population 25.26% vs 2.6%.^34^

**Table 1.** Demographics and Hospital Stay Characteristics

| Table 1. Demographics and Hospital Stay Characteristics |  |  |  |
| --- | --- | --- | --- |
|  | Expired | Did Not Expire | All Patients |
| Age |  |  |  |
| Range | 18 - 90 | 18 - 90 | 18 - 90 |
| Average (SD) | 73.62 (13.54) | 50.58 (18.95) | 52.8 (19.71) |
| Gender |  |  |  |
| Female | 3625 (44.43%) | 40698 (53.22%) | 44323 (52.38%) |
| Male | 4534 (55.57%) | 35767 (46.78%) | 40301 (47.62%) |
| Race |  |  |  |
| Asian | 200 (2.45%) | 1548 (2.02%) | 1748 (2.07%) |
| Black | 1284 (15.74%) | 18326 (23.97%) | 19610 (23.17%) |
| Hispanic | 16 (0.20%) | 166 (0.22%) | 182 (0.22%) |
| Multiracial/Other | 1561 (19.13%) | 19812 (25.91%) | 21373 (25.26%) |
| Natives | 17 (0.21%) | 150 (0.20%) | 167 (0.20%) |
| White | 5081 (62.27%) | 36463 (47.69%) | 41544 (49.09%) |
| Length of Stay |  |  |  |
| Range | 0 - 170 | 0 - 196 | 0 - 196 |
| Average (SD) | 11.65 (10.19) | 3.36 (6.09) | 4.16 (7.04) |
| Discharge Category |  |  |  |
| Expired | 8159 (100.00%) | N/A | 8159 (9.64%) |
| Home | N/A | 67421 (88.17%) | 67421 (79.67%) |
| Hospital | N/A | 2813 (3.68%) | 2813 (3.32%) |
| Rehabilitation Facility | N/A | 6231 (8.15%) | 6231 (7.36%) |
| Payment Type |  |  |  |
| Charity | 15 (0.18%) | 427 (0.56%) | 442 (0.52%) |
| Government | 742 (9.09%) | 18932 (24.76%) | 19674 (23.25%) |
| Standard Insurance | 665 (8.15%) | 19608 (25.64%) | 20273 (23.96%) |
| Medicare/caid | 6508 (79.76%) | 31417 (41.09%) | 37925 (44.82%) |
| Other | 229 (2.81%) | 6081 (7.95%) | 6310 (7.46%) |
| Admission Source |  |  |  |
| From Home | 6612 (81.04%) | 72643 (95.00%) | 79255 (93.66%) |
| Hospital Transfer | 1547 (18.96%) | 3822 (5.00%) | 5369 (6.34%) |
| Encounter Type |  |  |  |
| Emergency | 185 (2.27%) | 38818 (50.77%) | 39003 (46.09%) |
| Inpatient | 7973 (97.72%) | 35072 (45.87%) | 43045 (50.87%) |
| Observation | 1 (0.01%) | 2084 (2.73%) | 2085 (2.46%) |
| Same Day Surgery | 0 (0%) | 491 (0.64%) | 491 (0.58%) |

Comorbidity summary statistics are presented in Table 2. The three most common comorbidities were Hypertension (n=39154, 46.27%), Diabetes (n=24352, 28.78%), Sepsis (n=17122, 20.23%). Table 3 stratifies age, insurance status, and comorbidities by race. The majority of our patients have some type of insurance with the most common types being medicare/medicaid (44.82%), private insurance (23.96%), and government insurance through the state or through the healthcare exchange (23.25%). There were very few patients with charity pay (0.52%) or other sources of pay that do not fall into the above categories (7.46%). Based on data from the Census Bureau whites are more likely to have private insurance but, in our population, the black population is more likely to have private insurance than whites (28.52% vs. 23.68%).^34^ Distribution of insurance types, and comorbidities between Black and White are similar as shown in Table 3.

**Table 2.** Comorbidities of Interest

|  | Expired | Did Not Expire | All Patients |
| --- | --- | --- | --- |
| Elixhauser Comorbidity Index |  |  |  |
| Range | 0 - 19 | 0 - 18 | 0 - 19 |
| Average (SD) | 2.92 (2.61) | 0.86 (1.63) | 1.06 (1.85) |
| Alcohol Abuse | 220 (2.70%) | 1016 (1.33%) | 1236 (1.46%) |
| Anxiety | 1308 (16.03%) | 4848 (6.34%) | 6156 (7.27%) |
| Bipolar Disorder | 137 (1.68%) | 743 (0.97%) | 880 (1.04%) |
| Cerebrovascular disease | 551 (6.75%) | 1109 (1.45%) | 1660 (1.96%) |
| Chronic Kidney Disease | 3119 (28.23%) | 6541 (8.55%) | 9660 (11.42%) |
| COPD | 890 (10.91%) | 2890 (3.78%) | 3780 (4.47%) |
| Depression | 1006 (12.33%) | 4254 (5.56%) | 5260 (6.22%) |
| Developmental disorders | 50 (0.61%) | 467 (0.61%) | 517 (0.61%) |
| Diabetes | 4227 (51.81%) | 20125 (26.32%) | 24352 (28.78%) |
| Heart Failure | 2651 (32.49%) | 5504 (7.20%) | 8155 (9.64%) |
| Hypertension | 6715 (82.30%) | 32439 (42.42%) | 39154 (46.27) |
| Hypotension | 651 (7.98%) | 1375 (1.80%) | 2026 (2.39%) |
| Long Term Opioid Use | 231 (2.83%) | 925 (1.21%) | 1156 (1.37%) |
| Myocardial Infarction | 1202 (14.73%) | 1268 (1.66%) | 2470 (2.92%) |
| Opioid Abuse | 79 (0.97%) | 255 (0.33%) | 334 (0.39%) |
| Pregnancy | 6 (0.07%) | 2598 (3.40%) | 2604 (3.08%) |
| Psychoses | 361 (4.42%) | 1579 (2.06%) | 1940 (2.29%) |
| PTSD | 65 (0.80%) | 430 (0.56%) | 495 (0.58%) |
| Pulmonary Embolism | 225 (2.76%) | 643 (0.84%) | 868 (1.03%) |
| Schizophrenia | 211 (2.59%) | 859 (1.12%) | 1070 (1.26%) |
| Sedative Abuse | 50 (0.61%) | 65 (0.09%) | 115 (0.14%) |
| Sepsis | 5564 (68.19%) | 11558 (15.12%) | 17122 (20.23%) |
| Stimulant Abuse | 44 (0.54%) | 354 (0.46%) | 398 (0.47%) |
| Tobacco Abuse | 423 (5.18%) | 5608 (7.33%) | 6031 (7.13%) |

**Table 3.** Payment Type, Age, and Comorbidities by Race

| Table 3 Payment Type, Age, and Comorbidities by Race |  |  |  |  |  |  |  |
| --- | --- | --- | --- | --- | --- | --- | --- |
|  | Asian<br>(N=1,748) | Black<br>(N=19,610) | Hispanic<br>(N=182) | Multiracial/Other<br>(N=21,373) | Natives<br>(N=167) | White<br>(N=41,544) | Total<br>(N=84,624) |
| Payment Type |  |  |  |  |  |  |  |
| Charity | 0 (0.00%) | 89 (0.45%) | 2<br>(1.10%) | 152 (0.71%) | 1<br>(0.60%) | 198<br>(0.48%) | 442<br>(0.52%) |
| Gov | 182<br>(10.41%) | 3,871<br>(19.74%) | 69<br>(37.91%) | 8,205 (38.39%) | 40<br>(23.95%) | 7,307<br>(17.59%) | 19,674<br>(23.25%) |
| Insurance | 598<br>(34.21%) | 5,593<br>(28.52%) | 33<br>(18.13%) | 4,177 (19.54%) | 36<br>(21.56%) | 9,836<br>(23.68%) | 20,273<br>(23.96%) |
| Medicare/caid | 769<br>(43.99%) | 8,929<br>(45.53%) | 67<br>(36.81%) | 7,110 (33.27%) | 86<br>(51.50%) | 20,964<br>(50.46%) | 37,925<br>(44.82%) |
| Other | 199<br>(11.38%) | 1,128<br>(5.75%) | 11<br>(6.04%) | 1,729 (8.09%) | 4<br>(2.40%) | 3,239<br>(7.80%) | 6,310<br>(7.46%) |
| Age |  |  |  |  |  |  |  |
| Median<br>(IQR) | 57 (43-71) | 48 (33-63) | 50 (37-62) | 47 (34-60) | 49 (36-65) | 58 (41-74) | 52 (36-68) |
| Range | 18-91 | 18-91 | 19-91 | 18-91 | 18-91 | 18-91 | 18-91 |
| Elixhauser<br>Comorbidity<br>Index |  |  |  |  |  |  |  |
| Median<br>(IQR) | 1 (0-2) | 1 (0-2) | 1 (0-2) | 0 (0-2) | 1 (0-2) | 1 (0-2) | 1 (0-2) |
| Range | 0-12 | 0-19 | 0-7 | 0-17 | 0-13 | 0-21 | 0-21 |
| Alcohol Abuse | 10<br>(0.57%) | 189<br>(0.96%) | 2<br>(1.10%) | 291 (1.36%) | 3<br>(1.80%) | 741<br>(1.78%) | 1,236<br>(1.46%) |
| Anxiety | 82<br>(4.69%) | 869<br>(4.43%) | 19<br>(10.44%) | 1,121 (5.24%) | 15<br>(8.98%) | 4,050<br>(9.75%) | 6,156<br>(7.27%) |
| Bipolar<br>Disorder | 5 (0.29%) | 188<br>(0.96%) | 1<br>(0.55%) | 97 (0.45%) | 3<br>(1.80%) | 586<br>(1.41%) | 880<br>(1.04%) |
| Cerebrovascular<br>disease | 32<br>(1.83%) | 310<br>(1.58%) | 1<br>(0.55%) | 222 (1.04%) | 6<br>(3.59%) | 1,089<br>(2.62%) | 1,660<br>(1.96%) |
| Depression | 92<br>(5.26%) | 764<br>(3.90%) | 8<br>(4.40%) | 709 (3.32%) | 12<br>(7.19%) | 3,675<br>(8.85%) | 5,260<br>(6.22%) |
| Developmental<br>disorders | 4 (0.23%) | 103<br>(0.53%) | 0<br>(0.00%) | 56 (0.26%) | 0<br>(0.00%) | 354<br>(0.85%) | 517<br>(0.61%) |
| Heart Failure | 138<br>(7.89%) | 1,902<br>(9.70%) | 11<br>(6.04%) | 1,144 (5.35%) | 23<br>(13.77%) | 4,937<br>(11.88%) | 8,155<br>(9.64%) |
| Hypotension | 53<br>(3.03%) | 383<br>(1.95%) | 1<br>(0.55%) | 368 (1.72%) | 6<br>(3.59%) | 1,215<br>(2.92%) | 2,026<br>(2.39%) |
| Long Term<br>Opioid Use | 14<br>(0.80%) | 163<br>(0.83%) | 1<br>(0.55%) | 275 (1.29%) | 4<br>(2.40%) | 699<br>(1.68%) | 1,156<br>(1.37%) |
| Myocardial | 57 | 459 | 3 | 486 (2.27%) | 8 | 1,457 | 2,470 |
| Infarction | (3.26%) | (2.34%) | (1.65%) |  | (4.79%) | (3.51%) | (2.92%) |
| Opioid Abuse | 3 (0.17%) | 48 (0.24%) | 0 (0.00%) | 45 (0.21%) | 3 (1.80%) | 235 (0.57%) | 334 (0.39%) |
| Pregnancy | 38 (2.17%) | 544 (2.77%) | 7 (3.85%) | 953 (4.46%) | 3 (1.80%) | 1,059 (2.55%) | 2,604 (3.08%) |
| Psychoses | 14 (0.80%) | 476 (2.43%) | 4 (2.20%) | 237 (1.11%) | 4 (2.40%) | 1,205 (2.90%) | 1,940 (2.29%) |
| PTSD | 4 (0.23%) | 86 (0.44%) | 2 (1.10%) | 84 (0.39%) | 1 (0.60%) | 318 (0.77%) | 495 (0.58%) |
| Pulmonary Embolism | 18 (1.03%) | 259 (1.32%) | 1 (0.55%) | 132 (0.62%) | 3 (1.80%) | 455 (1.10%) | 868 (1.03%) |
| Schizophrenia | 11 (0.63%) | 304 (1.55%) | 2 (1.10%) | 149 (0.70%) | 2 (1.20%) | 602 (1.45%) | 1,070 (1.26%) |
| Sedative Abuse | 5 (0.29%) | 12 (0.06%) | 0 (0.00%) | 29 (0.14%) | 1 (0.60%) | 68 (0.16%) | 115 (0.14%) |
| Sepsis | 467 (26.72%) | 3,428 (17.48%) | 32 (17.58%) | 4,210 (19.70%) | 34 (20.36%) | 8,951 (21.55%) | 17,122 (20.23%) |
| Stimulant Abuse | 5 (0.29%) | 108 (0.55%) | 0 (0.00%) | 91 (0.43%) | 0 (0.00%) | 194 (0.47%) | 398 (0.47%) |
| Tobacco Abuse | 70 (4.00%) | 1,650 (8.41%) | 9 (4.95%) | 1,275 (5.97%) | 22 (13.17%) | 3,005 (7.23%) | 6,031 (7.13%) |

Outcome variables of interest were LoS and mortality. Table 4 includes negative binomial regression with independent variables of age, insurance, sex, comorbidites with LoS as the dependent variable. The results are shown as incidence rate ratios, interpretable as would be odds ratios. Table 5 contains a logistic regression of in-hospital mortality data, using age, insurance status, sex, comorbidities as the dependent variables.

**Table 4.**
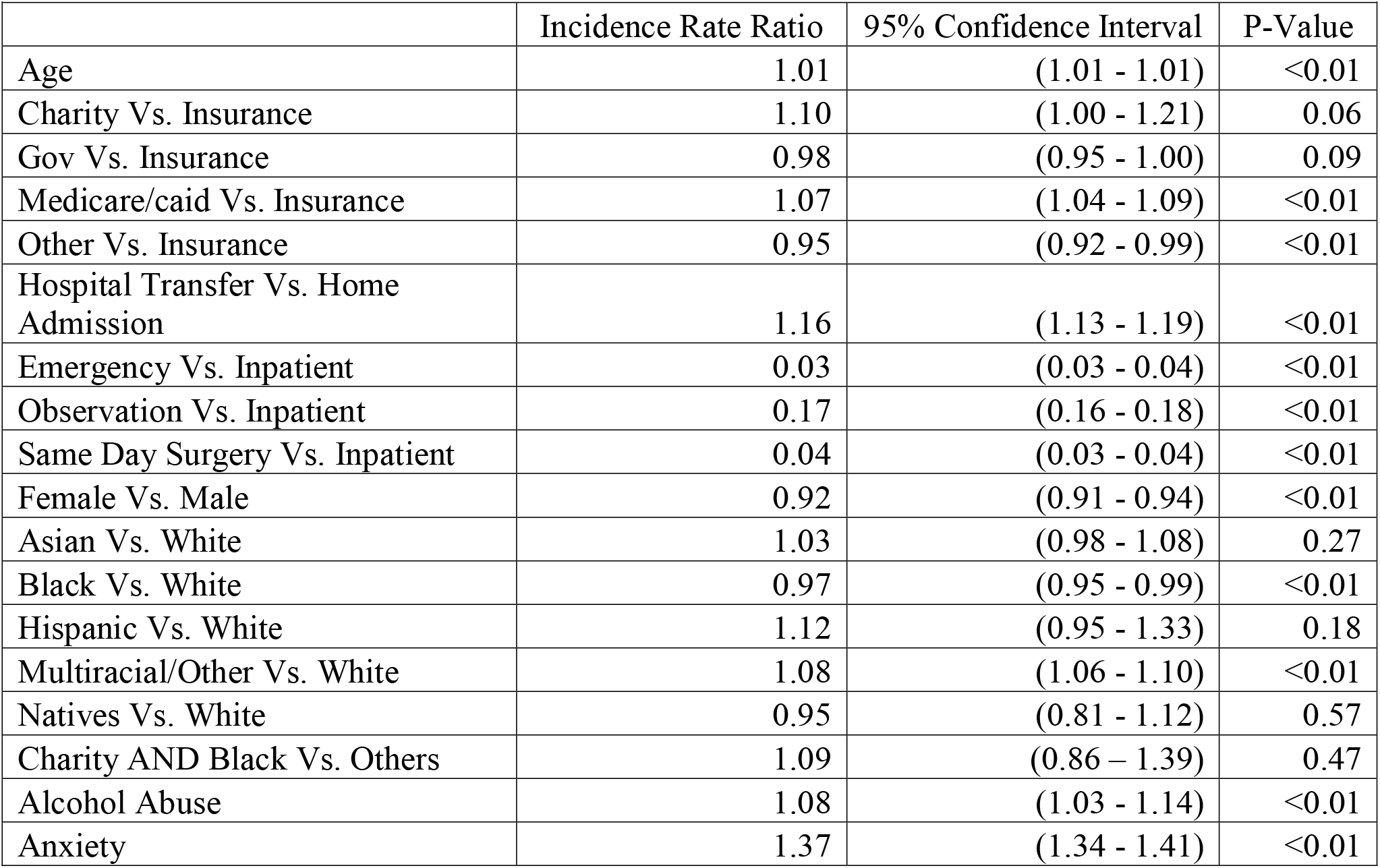
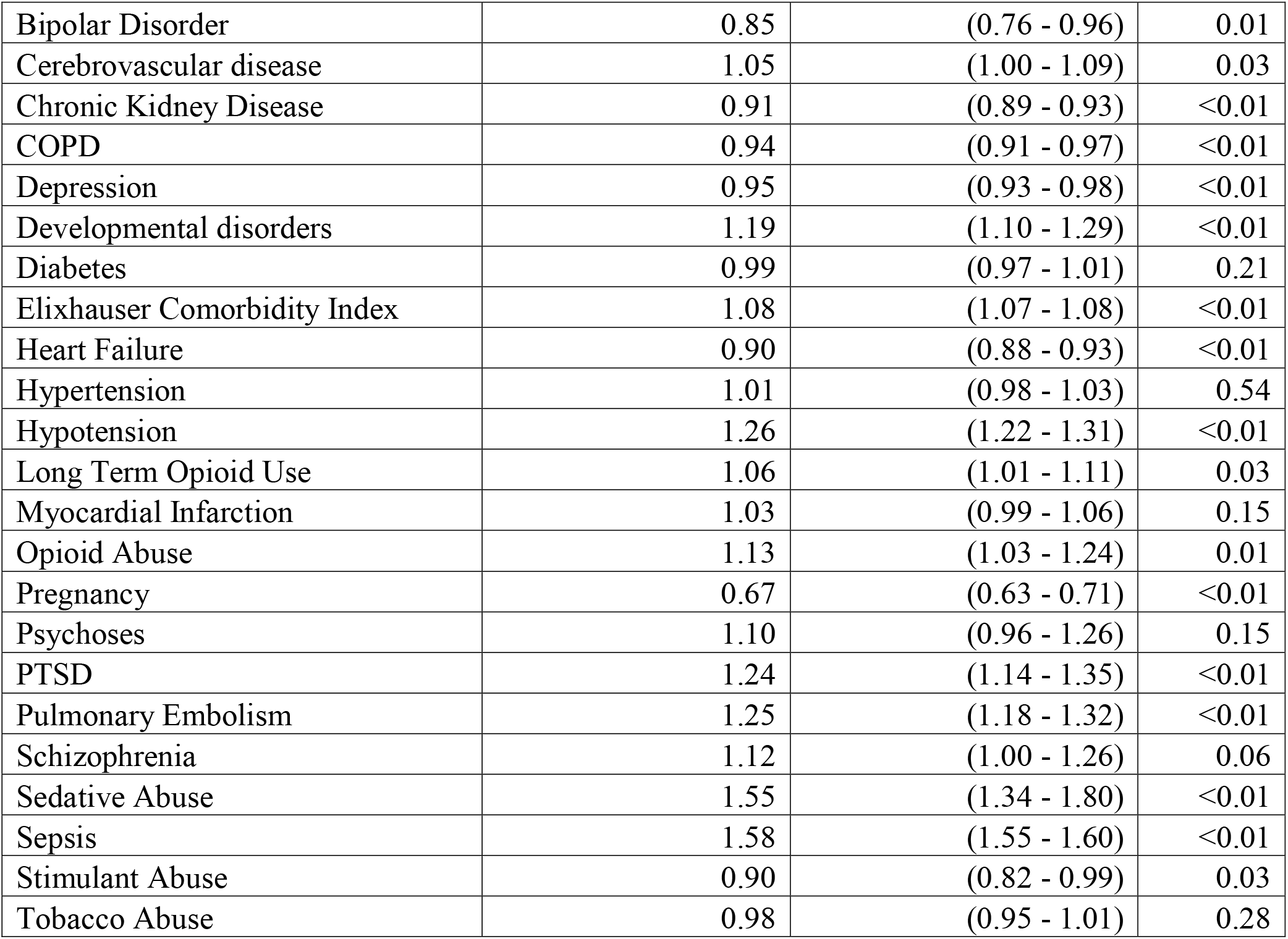
Negative Binomial Regression – Length of Stay (in Days)

| Table 4. Negative Binomial Regression – Length of Stay (in Days) |  |  |  |
| --- | --- | --- | --- |
|  | Incidence Rate Ratio | 95% Confidence Interval | P-Value |
| Age | 1.01 | (1.01 - 1.01) | <0.01 |
| Charity Vs. Insurance | 1.10 | (1.00 - 1.21) | 0.06 |
| Gov Vs. Insurance | 0.98 | (0.95 - 1.00) | 0.09 |
| Medicare/caid Vs. Insurance | 1.07 | (1.04 - 1.09) | <0.01 |
| Other Vs. Insurance | 0.95 | (0.92 - 0.99) | <0.01 |
| Hospital Transfer Vs. Home Admission | 1.16 | (1.13 - 1.19) | <0.01 |
| Emergency Vs. Inpatient | 0.03 | (0.03 - 0.04) | <0.01 |
| Observation Vs. Inpatient | 0.17 | (0.16 - 0.18) | <0.01 |
| Same Day Surgery Vs. Inpatient | 0.04 | (0.03 - 0.04) | <0.01 |
| Female Vs. Male | 0.92 | (0.91 - 0.94) | <0.01 |
| Asian Vs. White | 1.03 | (0.98 - 1.08) | 0.27 |
| Black Vs. White | 0.97 | (0.95 - 0.99) | <0.01 |
| Hispanic Vs. White | 1.12 | (0.95 - 1.33) | 0.18 |
| Multiracial/Other Vs. White | 1.08 | (1.06 - 1.10) | <0.01 |
| Natives Vs. White | 0.95 | (0.81 - 1.12) | 0.57 |
| Charity AND Black Vs. Others | 1.09 | (0.86 – 1.39) | 0.47 |
| Alcohol Abuse | 1.08 | (1.03 - 1.14) | <0.01 |
| Anxiety | 1.37 | (1.34 - 1.41) | <0.01 |
| Bipolar Disorder | 0.85 | (0.76 - 0.96) | 0.01 |
| Cerebrovascular disease | 1.05 | (1.00 - 1.09) | 0.03 |
| Chronic Kidney Disease | 0.91 | (0.89 - 0.93) | <0.01 |
| COPD | 0.94 | (0.91 - 0.97) | <0.01 |
| Depression | 0.95 | (0.93 - 0.98) | <0.01 |
| Developmental disorders | 1.19 | (1.10 - 1.29) | <0.01 |
| Diabetes | 0.99 | (0.97 - 1.01) | 0.21 |
| Elixhauser Comorbidity Index | 1.08 | (1.07 - 1.08) | <0.01 |
| Heart Failure | 0.90 | (0.88 - 0.93) | <0.01 |
| Hypertension | 1.01 | (0.98 - 1.03) | 0.54 |
| Hypotension | 1.26 | (1.22 - 1.31) | <0.01 |
| Long Term Opioid Use | 1.06 | (1.01 - 1.11) | 0.03 |
| Myocardial Infarction | 1.03 | (0.99 - 1.06) | 0.15 |
| Opioid Abuse | 1.13 | (1.03 - 1.24) | 0.01 |
| Pregnancy | 0.67 | (0.63 - 0.71) | <0.01 |
| Psychoses | 1.10 | (0.96 - 1.26) | 0.15 |
| PTSD | 1.24 | (1.14 - 1.35) | <0.01 |
| Pulmonary Embolism | 1.25 | (1.18 - 1.32) | <0.01 |
| Schizophrenia | 1.12 | (1.00 - 1.26) | 0.06 |
| Sedative Abuse | 1.55 | (1.34 - 1.80) | <0.01 |
| Sepsis | 1.58 | (1.55 - 1.60) | <0.01 |
| Stimulant Abuse | 0.90 | (0.82 - 0.99) | 0.03 |
| Tobacco Abuse | 0.98 | (0.95 - 1.01) | 0.28 |

**Table 5.** Logistic Regression – In Hospital Mortality

|  | Odds Ratio | 95% Confidence Interval |
| --- | --- | --- |
| Age | 1.06 | (1.06 - 1.07) |
| Charity Vs. Insurance | 0.99 | (0.54 - 1.69) |
| Gov Vs. Insurance | 1.55 | (1.37 - 1.75) |
| Medicare/caid Vs. Insurance | 1.11 | (1.00 - 1.23) |
| Other Vs. Insurance | 1.14 | (0.96 - 1.35) |
| Hospital Transfer Vs. Home Admission | 1.76 | (1.63 - 1.91) |
| Emergency Vs. Inpatient | 0.21 | (0.18 - 0.25) |
| Observation Vs. Inpatient | 0.02 | (0.00 - 0.07) |
| Same Day Surgery Vs. Inpatient | 0.03 | (0.01 - 0.17) |
| Female Vs. Male | 0.84 | (0.79 - 0.89) |
| Asian Vs. White | 0.92 | (0.76 - 1.10) |
| Black Vs. White | 0.81 | (0.75 - 0.87) |
| Hispanic Vs. White | 1.64 | (0.87 - 2.92) |
| Multiracial/Other Vs. White | 0.99 | (0.92 - 1.07) |
| Natives Vs. White | 1.10 | (0.58 - 2.00) |
| Charity AND Black vs Others | 1.00 | (0.93 – 1.07) |
| Alcohol Abuse | 1.15 | (0.96 - 1.38) |
| Anxiety | 1.37 | (1.25 - 1.49) |
| Bipolar Disorder | 0.98 | (0.64 - 1.48) |
| Cerebrovascular disease | 1.13 | (1.00 - 1.29) |
| Chronic Kidney Disease | 1.08 | (1.04 - 1.12) |
| COPD | 0.98 | (0.94 - 1.02) |
| Depression | 0.77 | (0.70 - 0.85) |
| Developmental disorders | 1.12 | (0.79 - 1.54) |
| Diabetes | 0.96 | (0.94 - 0.99) |
| Elixhauser Comorbidity Index | 1.14 | (1.13 - 1.16) |
| Heart Failure | 1.04 | (0.95 - 1.13) |
| Hypertension | 0.94 | (0.92 - 0.97) |
| Hypotension | 1.53 | (1.36 - 1.72) |
| Long Term Opioid Use | 0.99 | (0.84 - 1.18) |
| Myocardial Infarction | 2.50 | (2.27 - 2.77) |
| Opioid Abuse | 1.51 | (1.10 - 2.06) |
| Pregnancy | 0.35 | (0.10 - 0.88) |
| Psychoses | 0.80 | (0.50 - 1.29) |
| PTSD | 0.74 | (0.54 - 1.00) |
| Pulmonary Embolism | 1.46 | (1.21 - 1.76) |
| Schizophrenia | 1.13 | (0.73 - 1.73) |
| Sedative Abuse | 2.19 | (1.39 - 3.45) |
| Sepsis | 4.86 | (4.58 - 5.17) |
| Stimulant Abuse | 1.30 | (0.87 - 1.90) |
| Tobacco Abuse | 0.86 | (0.75 - 0.97) |

In our population, the results of negative binomial regression found blacks had a decreased length of stay (IRR 0.97, CI 0.95-0.99) and decreased mortality (OR 0.81, CI 0.75-0.97). Though, the difference in magnitude for length of stay was quite small. For every increase in age by one year, the length of stay is increased by 1.01 times (or 1%) which compounds in older individuals. Increasing age also resulted in increased mortality (OR 1.06, CI 1.06-1.07). Medicare and Medicaid patients were not admitted as long as those with private insurance (IRR 1.07, CI 1.04-1.09) though again the magnitude of difference was small. On the other hand Medicare and Medicaid patients had higher odds of in-hospital mortality than those with private insurance (OR 1.11, CI 1.00-1.23). Females had shorter hospital stay (IRR 0.92 CI 0.91-0.94) and had significantly lower in-hospital mortality than males (OR 0.84 CI 0.79-0.89). Patients with anxiety had significantly increased admission lengths (IRR 1.37, CI 1.34-1.41) with increased in-hospital mortality (OR 1.37, CI 1.25-1.49). Length of stay and in-hospital mortality were also increased in patients with cerebrovascular disease, hypotension, myocardial infarction, sepsis and pulmonary embolism as can be seen in tables 2 and 3. Patients with diagnosis of sedative abuse also had increased length of stay (IRR 1.55, CI 1.34 - 1.80) and a high in-hospital mortality (OR 2.19, CI 1.39-3.45). We also included the Elixhauser Comorbidity index and found length of stay (IRR 1.08, CI 1.07 - 1.08) and mortality increased (OR 1.14, CI 1.13-1.16) for each additional point on the scale.

## Discussion

We quantitatively assessed the relationship between several SDoH and comorbidities in a COVID-19 cohort to evaluate their impact on LoS and in-hospital mortality. When using insurance type as a proxy for social determinants we found that when the population of interest has a more equivalent distribution of socioeconomic status as well as comorbidities, the minority groups fared the same or better in terms of LOS and mortality as compared to whites.

Our black population is more likely to have private insurance than whites (28.52% vs. 23.68). In our population, blacks as compared to whites had a decreased LOS P<0.01 and decreased mortality (OR 0.81. CI 0.75-0.97). The white population was found to have higher rates of heart disease but equivalent elixhauser comorbidity indices. This may be the result of the fact that HCA serves a wealthier population in higher income zip codes. By using insurance status as a proxy to measure SDoH differences, we found that when minority groups are in a similar environment they perform equally well or better to whites in terms of length of stay and mortality in COVID-19 infections. This would indicate that genetic differences between races likely are not the cause of racial disparities in COVID-19 but the incidence of comorbidities and other social determinants of health like insurance status are likely the responsible factors for these differences seen in other studies as noted previously.

We have shown age, sepsis, hypotension on arrival to the hospital as well as other comorbid conditions like cerebrovascular disease, myocardial infarction are associated with increased LOS. Further, the elixhauser comorbidity index demonstrates that an increase in comorbidities is a very important factor in predicting LoS and mortality in patients with COVID-19. Our data supplements much of the data cited and published by the CDC.^35^ Timely identification of these comorbidities will be helpful in triaging patients that arrive at the hospital and are also significant predictors of mortality outside the setting of COVID-19. Our data also indicates a high success rate of HCA hospitals in their ability to triage patients to the proper setting based on length of stay and mortality.

In patients with anxiety the LoS and mortality increased significantly. We hypothesize that this may be due to difficulty with oxygenation modalities (e.g., BIPAP, CPAP, etc) that require a facemask and can often exacerbate anxiety. Very little research has been done that has shown the effects of anxiety in BIPAP and determining appropriate sedation management for these patients.^36^ This fascinating finding sets the stage for further research into mechanisms of oxygenation in these groups for better outcomes. Opioid and sedative abuse also increased mortality and length of stay. The etiology of this finding requires further research.

Although we have a more robust dataset than many previous studies, there are several limitations to this study. Our population of interest is different from prior work, thus generalization of these findings may not be appropriate. Applying our population’s findings with a more uniform wealth distribution and different distribution of commodities to a population that looks more like the US may result in inappropriate conclusions and triaging of patients. Also, given the timeframe of when our data was obtained, some hospitals within HCA may have not had as robust of a testing infrastructure, and some patient subtypes may not have been included. Additionally, there are other SDoH and confounding variables that are not included in our data, and our use of insurance status to reflect socioeconomic status is not as well validated as using a patient’s zip code. Finally, our data does not include patients that died outside of the hospital. Certain groups without access to healthcare were likely not included in our population. Not including these patients likely took some extremely sick and low resource patients out of our analysis. Further research may look at the effects on these outcomes after the introduction of vaccines and how that has affected the distribution of outcomes.

## Conclusion

Using the large HCA cohort, we found that when black patients have higher rates of private insurance, they have shorter hospitalizations and lower mortality than whites, when diagnosed with Covid-19. This supports the fact that Covid-19 racial differences are most likely the result of differences in socio-economic status.

This research was supported (in whole or in part) by HCA Healthcare and/or an HCA Healthcare affiliated entity. The views expressed in this publication represent those of the author(s) and do not necessarily represent the official views of HCA Healthcare or any of its affiliated entities.

## Data Availability

Data is property of HCA healthcare and will not be shared in it's original form.

